# A model-based evaluation of the efficacy of COVID-19 social distancing, testing and hospital triage policies

**DOI:** 10.1101/2020.04.20.20073213

**Authors:** Audrey McCombs, Claus Kadelka

## Abstract

We present a stochastic compartmental network model of SARS-CoV-2 and COVID-19 exploring the effects of policy choices in three domains: social distancing, hospital triaging, and testing. We distinguished between high-risk and low-risk members of the population, and modeled differences in social interactions due to context, risk level, infection status, and testing status. The model incorporates many of the currently important characteristics of the disease, including overcapacity in the healthcare system and uncertainties surrounding the proportion and transmission potential of asymptomatic cases. We compared current policy guidelines from public health agencies with alternative options, and investigated the effects of policy decisions on the overall proportion of COVID-19-related deaths. Our results support current policies to contain the outbreak but also suggest possible refinements, including emphasizing the need to reduce public, random contacts more than private contacts, and testing low-risk symptomatic individuals before high-risk symptomatic individuals. Our model furthermore points to interactions among the three policy domains; the efficacy of a particular policy choice depends on other implemented policies. Finally, our results provide an explanation for why societies like Germany, with lower average rates of social contact, are more successful at containing the outbreak than highly social societies such as Italy, despite the implementation of similar policy measures.

## 1 Introduction

On December 31, 2019, a pneumonia of unknown cause was reported to the World Health Organization (WHO) Country Office in China. The WHO named the disease COVID-19, caused by the novel coronavirus SARS-CoV-2, and declared the outbreak a pandemic on March 11, 2020. As of the time of this writing, April 19, 2020, the WHO has reported 2,241,359 confirmed cases and 152,551 deaths in 210 countries [1]. There is currently no vaccine for SARS-CoV-2 and no antiviral medication that specifically targets the virus. In response to the pandemic, public health organizations have deployed plans developed to respond to a possible pandemic similar to the influenza outbreak of 1918, which killed an estimated 50 million people world-wide [2]. These plans describe, among other things, policies related to social distancing and the allocation of scarce healthcare resources. Policy guidelines for testing individuals for exposure to SARS-CoV-2 are less well-developed, and a shortage of available tests and testing facilities has hampered response efforts in many countries.

The spread of an infectious disease can be strongly influenced by human behavior [3–9]. One study estimates that three to four months of moderate social distancing could save 1.7 million lives in the United States by October 1, 2020 [10]. In an examination of different non-pharmaceutical interventions, other researchers found that a combination of case isolation, home quarantine, and social distancing of high-risk individuals could halve the number of deaths in the United State and Great Britain [3]. Another recent study projects that prolonged or intermittent social distancing may be necessary into 2022 in order to avoid exceeding hospital capacity thresholds [11]. Because the effects of COVID-19 seem dependent on demographics such as age, allowing for differences in social interaction behaviors due to demography is an important aspect of modeling this disease.

A defining characteristic of the current COVID-19 pandemic is a shortage of healthcare resources. Individual hospitals as well as local, state, and federal agencies have developed guidelines to help healthcare workers decide which patients will receive scarce, life-saving resources such as ventilators [12–16]. Guidelines are based on ethical principles and societal norms, and governing principles generally include the duty to care and the duty to treat people fairly. Strategies for determining who will receive scarce resources include: 1) first-come first-served, 2) randomized allocation (e.g., lottery), and 3) clinical judgment [12–14]. The purpose of these guidelines is to relieve individual clinicians of the burden of deciding on-the-spot how to allocate resources.

Public health officials must balance the needs for accessibility and accuracy when determining testing policy during a pandemic [17]. False positives during the initial stages of the pandemic and false negatives during the later phases can lead to biased estimates of infection prevalence and dynamics [18]. Guidelines from the Centers for Disease Control and Prevention (CDC) currently prioritize testing for hospitalized patients and healthcare facility workers with symptoms [19]. Individuals with second priority include high-risk individuals and first responders with symptoms, while third priority goes to other individuals with symptoms as well as health care workers and first responders. Current guidelines recommend testing people without symptoms only when the testing needs of higher-priority individuals have been met.

While many models examine the effects of a single policy domain on the dynamics of an infectious disease, e.g., [3, 5, 15, 20–23], to our knowledge there are no studies examining several policy domains simultaneously. Considering policy domains together can provide crucial insight into how different policy decisions interact. For example, a mandatory quarantine of symptomatic individuals combined with an efficient testing regime may reduce the total number of deaths more than would be expected if the effects of these policies were modeled individually. Policies can also interfere with each other, so that the combined effect is weaker than expected. Our model provides a tool for investigating these interaction effects among three policy domains that have received recent attention from public officials and the media: social distancing, hospital triaging, and testing.

Classical compartmental differential equation models are an invaluable tool for understanding the general course of an infectious disease at a population level. However, these models assume that any two individuals interact with equal probability (homogeneous mixing), which is not the case in real physical interaction networks and can result in significantly different disease dynamics [24–26]. A further simplifying assumption frequently made in compartmental models is that transition rates are constant (Poisson assumption), implying Markovian memorylessness and exponentially distributed transition times [27]. For COVID-19, this is clearly not the case [28, 29]. Our study avoids both these pitfalls by implementing a non-Markovian compartmental disease model evaluated on an interaction network, adopting a flexible modeling framework for more realistic disease dynamics [30].

Our stochastic compartmental network model simulates how the SARS-CoV-2 virus spreads through an abstract community of 1000 individuals. A 2-layer interaction network represents private and public social contacts as small-world and fully-connected graphs, respectively (Fig. 1a). Upon infection, susceptibles (S) transition through contagious compartments (exposed (E), asymptomatic (A), infected (I) and hospitalized (H)), finally resulting in death (D) or recovery (R) (Fig. 1b). Our model incorporates important characteristics of the current COVID-19 outbreak such as early transmissibility of the virus, asymptomatic cases (Fig. 1c), and age-dependent differential risk. We modeled behavioral differences associated with risk level and infection status, as well as the reduction in care caused by hospitals operating beyond their capacity thresholds (Fig. 1d). Our model also incorporates the uncertainty surrounding key epidemiological parameters such as the proportion and transmissibility of asymptomatic cases [31].

**Figure 1:**
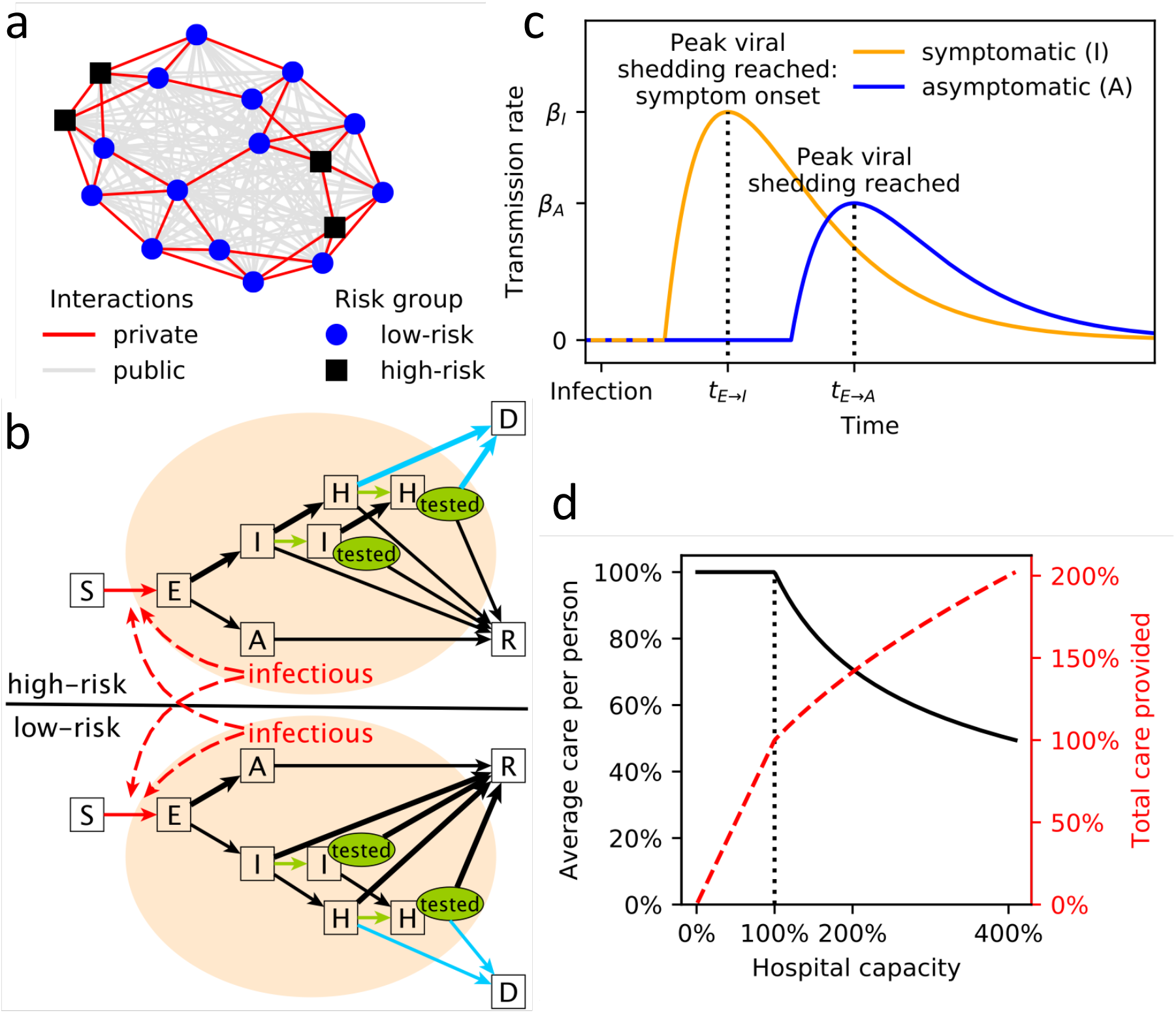
(a) Example of the two-layer interaction network used in this study. The private, small-world network (red edges) is shown on top of the public, fully-connected network (gray edges). Low-risk (blue circles) and high-risk (black squares) individuals are distinguished. (b) Illustration of the stochastic transmission model with compartments S = susceptible, E = exposed, A = asymptomatic, I = symptomatic, H = hospitalized, R = recovered, D = deceased. Individuals in I and H may received a positive test (green “tested” oval). Edges that are influenced by policy decisions are colored: red = social distancing, green = testing, blue = hospital triage. Branching probabilities at E, I and H are risk-group dependent and the edge of the respectively more likely transition is thicker. (c) Illustration of the time-dependent transmission rate of an exposed individual increasing until peak viral shedding, which coincides with transition to compartment I (if symptomatic) or A (otherwise). (d) Average care per person (blue solid line) and total care provided n(red dashed line) by a health care system with a capacity threshold of 100% operating at a certain level of (over)capacity. Once the capacity threshold is reached, the average care per person is 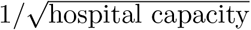.

The primary goal of this study was to evaluate the effects of various policies regarding social distancing, triaging, and testing on the disease dynamics of COVID-19. The abstract nature of our model enables a reliable evaluation of the relative, qualitative efficacy of different policy decisions in reducing COVID-19-related mortality, despite current uncertainty in various key parameters. The model can be easily updated and expanded once more accurate parameter estimates are available, and can be tailored to a specific community or country in order to evaluate the quantitative effects of policies being considered for implementation.

## 2 Methods

### 2.1 Model description

We modeled the spread of an imported case of SARS-CoV-2 across a multi-layered physical interaction network of size *N* = 1000 as an abstract proxy for a town or local community. We considered a closed population, and given the short time frame also chose not to include birth, deaths, or migration events. We distinguished between two types of interactions: First, private (local) interactions (e.g., friends, school and work colleagues) were modeled using a Watts-Strogatz small-world network with average connectivity of *k* neighbors and 5% probability of edge rewiring (Fig. 1a). (Edge rewiring refers to the replacement of edges in a regular network according to a stochastic algorithm so that properties of small-world networks, such as clustering and short path lengths, are achieved [32].) Second, public (global) interactions (e.g., grocery shopping, banking) were modeled using a fully connected network [33]. We assumed that, in the absence of an epidemic, an individual has on average the same number of *k* private and *k* public interactions, and thus assigned a weight of *k/*(*N−* 1) to each edge in the public interaction network. A multi-national study found, on average, between 8 and 20 per-person per-day contacts [9], so we considered *k∈ {*4, 6 (baseline), 10*}*.

We modeled the differential risk associated with COVID-19 by distinguishing between high-risk individuals (older individuals or individuals with known comorbidities [34]) and low-risk individuals (younger individuals without known comorbidities). Each node represented a high-risk (low-risk) individual with probability *p*^high-risk^ = 1*/*3 (*p*^low-risk^ = 2*/*3). Our model includes seven qualitatively different compartments: S = susceptible, E = exposed (infectivity increases), A = asymptomatic (low infectivity), I = symptomatic (high infectivity), H = requiring hospitalization due to severe infection (high infectivity), and two final compartments R = recovered (no infectivity) and D = died from COVID-19 infection (Fig. 1b). The length of time recovered individuals remain immune is currently unknown, however given the short time frame (weeks to months) of this model, we assumed no reinfections. To model the spread of an imported case of the virus in a fully susceptible community, we initialized the simulation with one random node in compartment E; all others started in S.

Model inputs related to the specific characteristics of SARS-CoV-2 and COVID-19 were derived from published literature where available (Table 1). Virus and disease parameters without established estimates were included as random variables from broad uniform distributions. We considered time to be discrete with one unit of time corresponding to a day. The lengths of time individuals spend in a contagious compartment were modeled as Poisson random variables with parameters derived from the literature (Table 2).

**Table 1:** Model parameters with reported estimates and sample sizes (*n*) from the literature where applicable and available. The third column shows the value we used or the range we sampled uniformly. ^*\**^[43] considers only the U.S. adult population. Adding 0-17 year-olds with an assumed high-risk rate of 21.2% (the estimate for 18-59yr olds) and projected 2020 US census data [44] yields an overall high-risk estimate of around 1*/*3. ^*†*^ numbers derived from Table 1 in [3] and projected 2020 US census data [44]

| Parameter | Meaning | Value/Range | References |
| --- | --- | --- | --- |
| **Interaction network parameters** |  |  |  |
| N | population size | 1000 |  |
| k | average number of private/public interactions | 4,6 (default),10 | [9] $2k \in [7.95, 19.77]$ |
| $p^{small-world}$ | probability that an edge in the private small-world network is re-wired | 5% | |
| $p^{high-risk}$ | proportion high-risk individuals | 1/3* | [43] 37.6% (n=430,000) |
| **Virus and disease parameters** |  |  |  |
| $\beta_I$ | transmission rate of symptomatic individuals | [0.05, 0.4] | [29] 9.6% (n=1,286) |
| $\beta_A$ | transmission rate of asymptomatic individuals | $[0, \beta_I]$ | no data |
| $p_{E \rightarrow A}$ | proportion of asymptomatic infections | [5%, 50%] | [37] 20.6-39.9% (n=634) |
| $\frac{p_{E \rightarrow A}^{low-risk}}{p_{E \rightarrow A}^{high-risk}}$ | ratio of asymptomatic infections in low-risk vs. high-risk individuals | [1, 5] | no data |
| $p_{I \rightarrow H}$ | probability of symptomatic individuals requiring hospitalization | 7% | [41] 5% (n=44,415)<br>[3, 42] 7.38%<sup>†</sup> |
| $\frac{p_{I \rightarrow H}^{high-risk}}{p_{I \rightarrow H}^{low-risk}}$ | ratio of high-risk vs. low-risk symptomatic individuals requiring hospitalization | [4, 10] | [3, 42] 6.47<sup>†</sup> |
| IFR | COVID-19 infection fatality rate (if hospitalized receive perfect care) | 1% | [42] 0.657% (n=44,672)<br>[3] 0.9%<sup>†</sup> |
| $\frac{p_{H \rightarrow D}^{high-risk}}{p_{H \rightarrow D}^{low-risk}}$ | ratio of high-risk vs. low-risk hospitalized individuals dying from COVID-19 | [4, 10] | [3, 42] 6.33<sup>†</sup> |

**Table 2:** Distributions used to model the time an individual spends in each transient compartment (all times in days). Mean (*μ*) and median (*m*) estimates and sample sizes (*n*) from the literature are reported.

| Parameter | Meaning | Distribution | Reported mean ($\mu$)/median ($m$) |
| --- | --- | --- | --- |
| $t_{E \rightarrow I}$ | time in exposed compartment if infection will be symptomatic | Poisson(5) | [28] $m=4$ (n=1,099)<br>[29] $\mu=5.95$, $m=4.8$ (n=183)<br>[45] $m=5.1$ (n=181)<br>[7] $\mu=4.2$ (n=140)<br>[46] $\mu=5.2$ (n=49) |
| $t_{E \rightarrow A}$ | time in exposed compartment if infection will be asymptomatic | Poisson(5) | no data, assumed to be distributed as $t_{E \rightarrow I}$ |
| $t_{I \rightarrow H}$ | transition time from symptom onset to hospitalization | Poisson(8) | [29] $\mu=4.64$, $m=3.41$ (n=391)<br>[47] $m=11$ (n=191)<br>[48] $m=7$ (n=138)<br>[49] $m=7$ (n=41) |
| $t_{I \rightarrow R}$ | transition time from symptom onset to recovery (end of viral shedding) | Poisson(20) | [29] $m \in [17.5, 22.9]$ age-dependent (n=228)<br>[42] $\mu=24.7$ (n=165)<br>[47] $m=20$ (n=137) |
| $t_{A \rightarrow R}$ | transition time from full viral shedding to recovery | Poisson(20) | no data, assumed to be distributed as $t_{I \rightarrow R}$ |
| $t_{H \rightarrow R}$ | transition time from hospitalization to recovery | Poisson(12) | [28] $\mu=12.8$, $m=12$ (n=1,099)<br>[7] $\mu=11.5$ (n=140)<br>[47] $m=12$ (n=137)<br>[50] $\mu=17.4$ (n=21) |

Upon infection, individuals transition from one compartment to the next - until they recover or die - based on a stochastic process (Fig. 1b). Susceptible individuals become infected (and transition to the exposed compartment E) through contact with contagious individuals. Contrary to SARS [35], recent reports indicate that SARS-CoV-2 can be transmitted before the onset of symptoms and by asymptomatic cases [36–38]. To account for this early transmissibility in our model, individuals in compartments E, A, I and H may all transmit the virus, with transmission rates dependent on the time since infection. We assumed that exposed individuals (compartment E) become contagious 2 days before peak viral load, which coincides with symptom onset in symptomatic cases (i.e., the latent period is two days shorter than the incubation period). Transmission rates over time typically follow a Gamma distribution [39]. Based on preliminary data [40], we used a Gamma-distributed transmission rate with shape=2 and scale=2 (Fig. 1c). We further assumed that asymptomatic cases cannot be more contagious than symptomatic ones. SARS-CoV-2 transmission rates are currently not well understood [29], so we considered a range of values for the peak transmission rate of symptomatic cases (at symptom onset), *β*_*I*_ *∈ U* (0.05, 0.4) and a dependent range for asymptomatic ones, *β*_*A*_ *∈U* (0, *β*_*I*_).

Once exposed individuals reach peak infectivity they transition to the asymptomatic (A) or symptomatic (I) compartment. The proportion of asymptomatic COVID-19 infections is currently unknown; we therefore sampled the overall proportion of asymptomatic infections from a uniform distribution, 1 *p*_*E→I*_ = *p*_*E→A*_ *∼U* (0.05, 0.5), and further sampled the ratio of asymptomatic infections in low-risk versus high-risk individuals from another uniform distribution,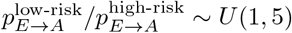.

A proportion of symptomatic individuals develop a severe infection requiring hospitalization; this rate may depend on the underlying health of the individual. We thus included two hospitalization parameters in the model (Table 1): *p*_*I→H*_ = 1 *− p*_*I→R*_ describes the overall proportion of symptomatic cases that eventually develop a severe infection and require hospitalization, while 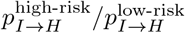 describes the increased likelihood of a high-risk individual requiring hospitalization. We fixed *p*_*I→H*_ = 7% and considered a range [4, 10] for the differential risk ratio [3, 41, 42].

We fit the overall proportion of hospitalized individuals dying from COVID-19, *p*_*H→D*_, to align with a COVID-19 infection fatality rate (IFR) of 1% [3, 42]. That is,

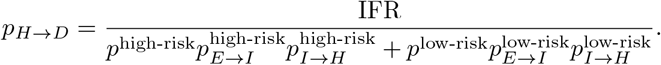

As before, we introduced a ratio describing the differential risk of dying from COVID-19 for high- and low-risk individuals,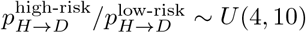. To simulate deaths in our network model, we made the simplifying assumption that each day a severely infected person has the same chance of dying from COVID-19 (i.e., the time to death is geometrically distributed with the distribution parameter corresponding to a per-day death rate). We fit the per-day death rate for low-risk and high-risk individuals to align with 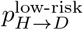 and 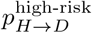, respectively.

Each day, the network model updates simultaneously as follows:

- Susceptibles may become infected through private or public interactions with contagious individuals. The interaction probabilities are based on the multi-layered interaction network.
- Newly infected individuals move to the compartment E and risk-group-dependent random variables are drawn describing the future course and transition times of the infection.
- Hospitalized individuals die at a risk-group-dependent per-day death rate.
- The transition times to the next compartment of all contagious individuals are reduced by a day. Individuals with a transition time of zero transition to the next compartment.

To investigate the effects of social distancing, triaging, and testing, we added additional features to this base stochastic network model, described in the next section.

### 2.2 Model extensions

#### 2.2.1 Social distancing

We modeled the general effects of social distancing policies with two parameters, private activity level *a*^*private*^ and public activity level *a*^*public*^, which describe the average degree to which an individual without symptoms (in compartments S, E, A or R) reduces private and public interactions. An individual who has not adopted social distancing behaviors has activity levels of 1, while perfect isolation corresponds to activity levels of 0.

We assumed that symptomatic individuals (in compartment I) further reduce their private and public activity levels due to symptoms and empathetic fear of infecting others at an average rate of *r*^symptoms^*∼ U* (0, 1). Similarly, we assumed that severely infected individuals requiring hospitalization (in compartment H) are completely isolated. Finally, we assumed that individuals in the high-risk group may, independently of their compartment, choose to reduce their activity levels more than the low-risk group. We therefore included an additional high-risk activity reduction, *r*^high-risk^*∼U* (0, 1).

The probability that two individuals who practice social distancing still meet, with the potential to infect one another, is given by a mass action-like product of their respective activity levels. For example, the probability that a symptomatic, low-risk individual meets a high-risk friend is given by *a*^*private*^(1*−r*^symptoms^) · *a*^*private*^(1*−r*^high-risk^).

In reality, each person decides individually how to adapt her social behavior in response to COVID-19. For this reason we assigned activity levels to an individual (node) rather than a contact (edge). To compare policy efficacies, however, we combined all individual-based activity levels into an overall, population-wide contact reduction rate. In a community without symptomatic infections (i.e., at the start of the simulation, before community members contract COVID-19), this overall contact reduction rate is a function of the underlying interaction network, the private and public activity levels, the additional activity reduction of high-risk individuals and the proportion of high-risk individuals.

#### 2.2.2 Hospital Triaging

The baseline model assumes unlimited healthcare resources, but in reality the number of hospital beds, ICU beds, ventilators, and trained health care professionals are all limited. We modeled limited healthcare resources by adding a capacity threshold under which the healthcare system can provide perfect care (3 beds for every 1000 individuals [51]). We assumed that this capacity threshold doubles during times of emergency, but the increase is not large enough to provide perfect care for all hospitalized individuals.

In the absence of data, we modeled the decrease in the average care provided per person with a square-root function, where average care per 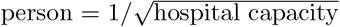. Once the number of individuals requiring hospitalization (in compartment H) rises above the capacity threshold, overall care is inadequate and triaging questions regarding resource allocation arise (Fig. 1d). We evaluated four options: (1) fill empty beds based on a wait list (corresponding to a first-come first-served strategy), (2) fill empty beds randomly (e.g., lottery), (3) fill empty beds with least-severely infected based on clinical judgment (in the model, clinical judgement corresponds to the known remaining time to recovery), and (4) provide the same level of imperfect care to each individual (e.g., sharing of a single ventilator among multiple patients). Under the first three policies, patients receiving care will receive perfect care until they recover or die, and the sole difference among these three policies is how empty beds are allocated.

In the baseline model, each hospitalized individual (in compartment H) who does not die on a given day moves closer to recovery. Under the imperfect care scenario, hospitalized individuals only move a partial day closer to recovery, corresponding to the amount of imperfect care they receive on that day. Individuals who receive perfect care move a full day closer to recovery, while individuals who receive only palliative care do not progress towards recovery.

#### 2.2.3 Testing

To evaluate the efficacy of testing policies in reducing COVID-19-related deaths, we assumed a fixed maximum number of tests available per day, and that testing begins as soon as the first person becomes symptomatic. We further assumed that, per CDC guidelines, severely infected individuals arriving at a hospital (compartment H) receive priority testing [52]. Remaining available tests are administered to symptomatic individuals (compartment I), and a shortage of tests precludes testing of individuals without symptoms (compartments S, E, A, and R). We compared two primary testing policies for symptomatic individuals: (i) test high-risk individuals first, or (ii) test low-risk individuals first. Further, within each primary testing policy, we compared two secondary testing policies: (i) test individuals in the order in which they developed symptoms (i.e., test first symptomatic first), or (ii) test individuals in the reverse order in which they developed symptoms (i.e., test recent symptomatic first). Finally, we included in the model a delay in test results of up to seven days.

While testing hospitalized individuals serves an essential clinical role, testing symptomatic individuals is solely preventive. Individuals who test positive are currently placed under quarantine, which in theory completely prevents virus transmission. In reality this is not always the case, especially when self-quarantine is conducted at home. We therefore included the average activity reduction of a positively tested individual as a further model parameter, and assumed a positive test yields a 80%*−*100% reduction in activity levels for the duration of the infection, in addition to the already-reduced activity levels due to symptoms, (i.e.,*r*^positive^ *∼ U* (0.8, 1)). For example, a high-risk, symptomatic individual who tested positive meets a low-risk friend with probability *a*^*private*^(1 *− r*^high-risk^)(1 *− r*^symptoms^)(1 *− r*^positive^) *· a*^*private*^.

### 2.3 Model analysis

Because the true values of many virus- and disease-related parameters are currently uncertain, we sampled all unknown parameters from a broad uniform distribution. For most analyses (Figs. 2,S5,3a,4a,b), we ran the model 10^6^ times, each time with a different parameter setting picked at random from the parameter space. To ensure sufficient coverage of the high-dimensional parameter space, for most analyses we opted for a large number of sample points versus replication. We sampled most parameters uniformly from their respective range. Only for the private and public activity levels as well as the additional high-risk contact reduction, we diverted from this approach and did not sample uniformly at random from [0, 1], in order to ensure that the distribution of overall contact reduction, which is derived from these three parameters and the underlying network, was wide enough to be representative (Table S1).

**Figure 2:**
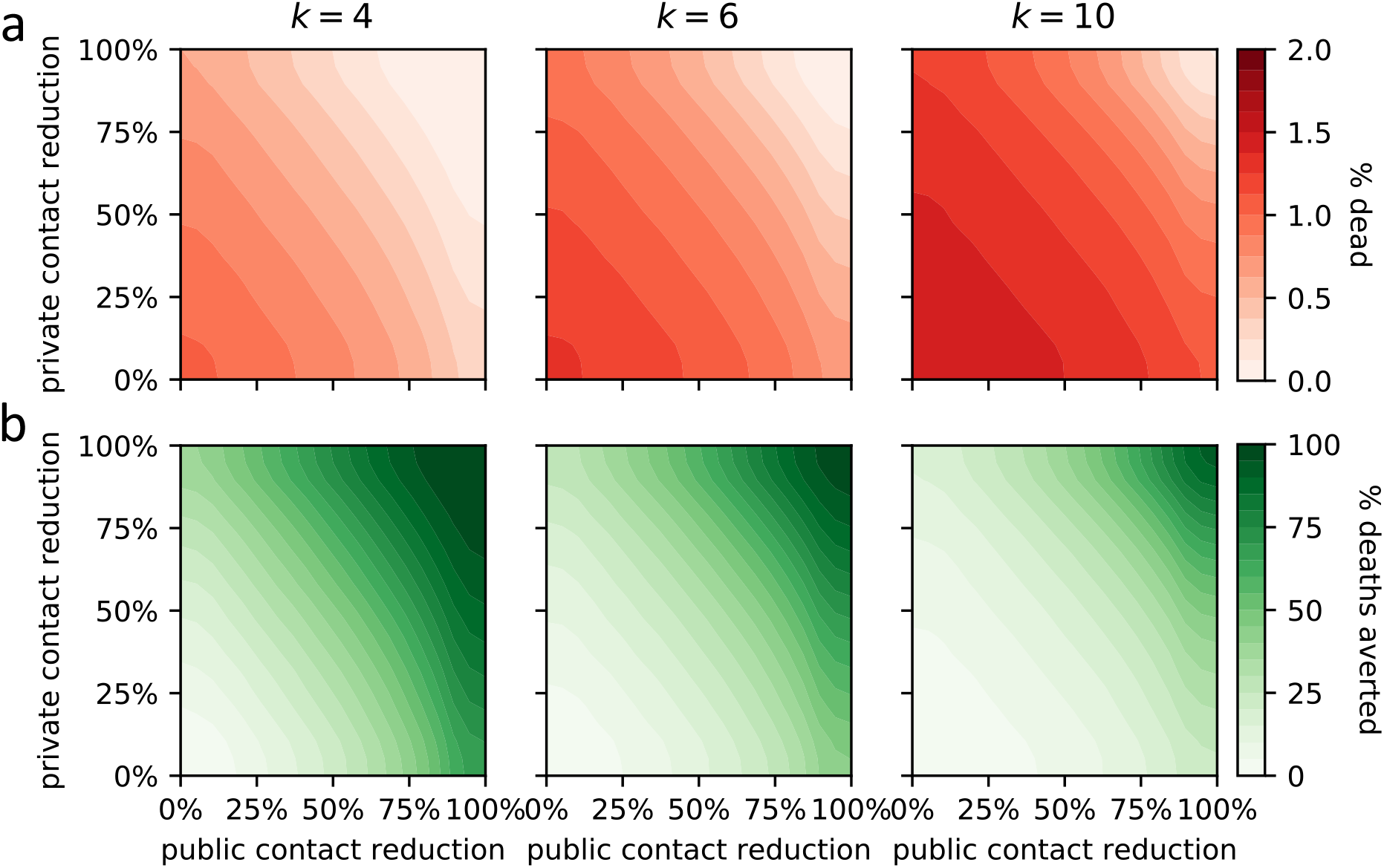
Effect of overall private and public contact reduction on the average proportion dead (red, top row) and the average percentage of averted deaths (green, bottom row). The latter is computed by comparison with no contact reduction. Results are shown for communities with *k* private and *k* public average contacts per day, for different values of *k*.

**Figure 3:**
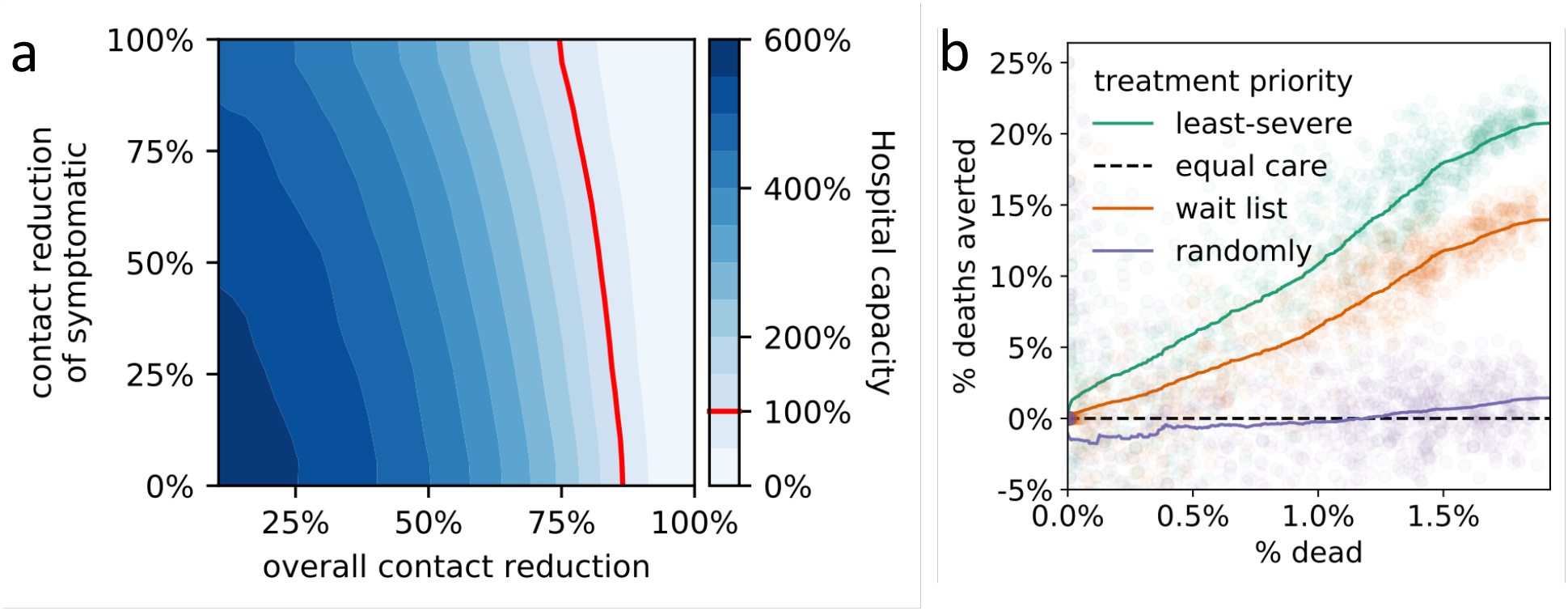
(a) Effect of overall contact reduction and additional contact reduction of symptomatic individuals on the projected peak hospital capacity. To the left of the hospital capacity threshold (red line), not all hospitalized individuals receive perfect care, resulting in longer recovery times and higher mortality. (b) Relative efficacy of four hospital triage policies for varying degrees of outbreak severity summarized by the average proportion of deaths (x-axis). Efficacies are computed by pairwise comparison of the projected death count with the imperfect-but-equal-care scenario.

When comparing the effect of triage policies we balanced coverage with precision by sampling 10^3^ parameter settings and running the model 250 times for each parameter setting for each triage policy. We initialized the runs of each triage policy with the same 250 random seeds to ensure our estimates were comparable. This approach works particularly well for the triage policies as triage choices do not affect the model until hospitals operate at overcapacity. To compare primary and secondary testing policies (Fig. 4c) we followed the same approach, except that we sampled from a lower-dimensional parameter space. We fixed the maximum number of tests per day at ten, assumed no delay, and considered only three and twenty levels of additional contact reduction by symptomatic individuals and high-risk individuals, respectively (details in second-to-last column of Table S1). Finally, when comparing the interaction between all policy domains (Fig. 5), we sampled 10^3^ parameter settings and ran the model 100 times for each parameter setting and each of the 2 *·*2 *·*4 = 16 combinations of policy choices (0 vs. 40 maximal tests per day; triage policies: imperfect but equal care vs. treat least severely infected first; four contact reduction levels: 0%, 25%, 50%, 75%; details in last column of Table S1).

**Figure 4:**
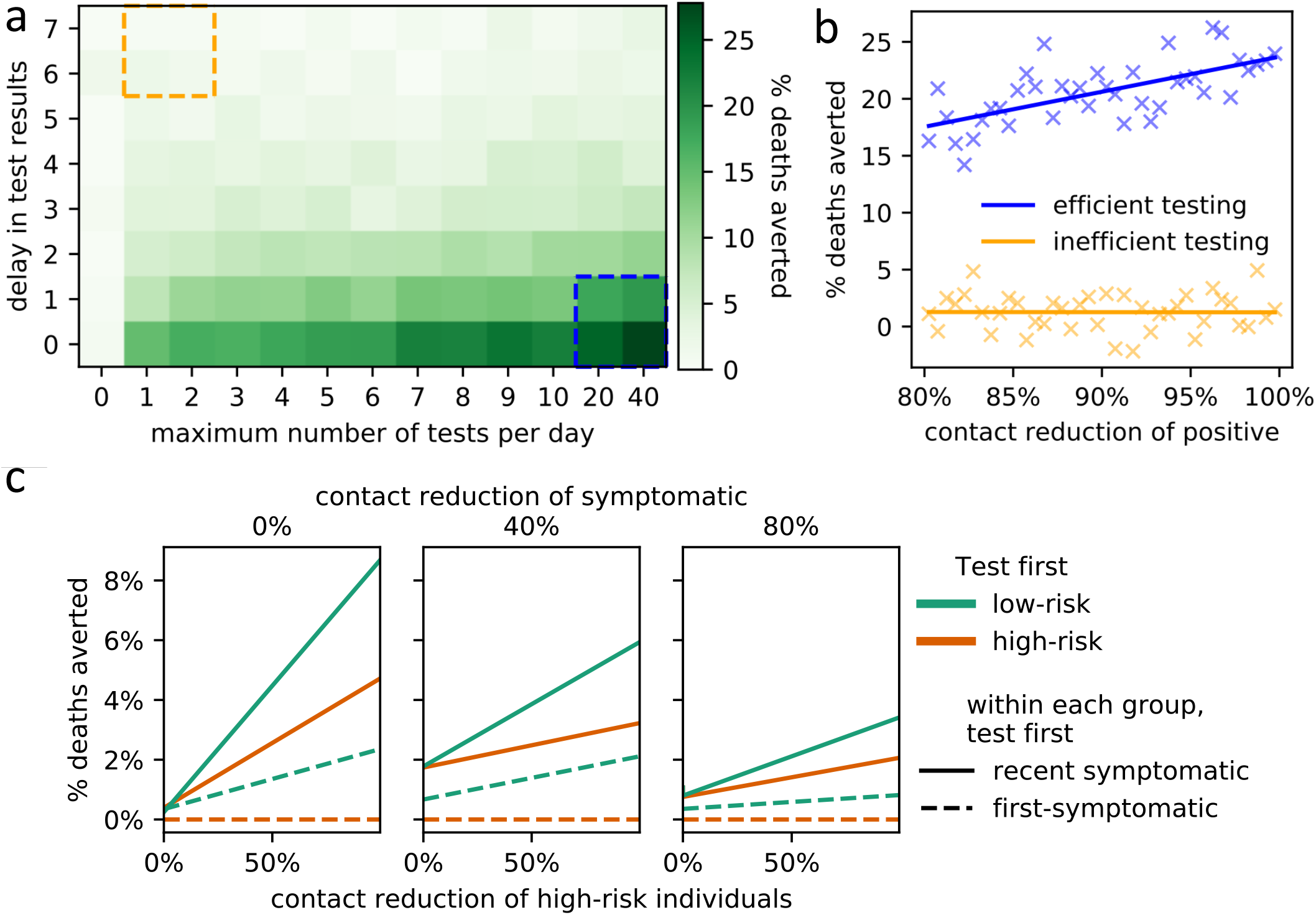
(a) Effect of increased numbers of tests (x-axis) and processing delays (y-axis) on the average percentage of averted deaths (compared to no testing; left-most column). (b) Effect (linear regression) of the average contact reduction by individuals with positive test results on the percentage of averted deaths under an efficient testing scenario (blue box lower right, panel a) and an inefficient testing scenario (orange box upper left, panel a). (c) Impact of policies regarding testing prioritization of symptomatic individuals on the average percentage of averted deaths (compared to the worst policy). The primary policy decision involves which risk group to prioritize (low-risk (green) or high-risk (orange)). The secondary policy decision involves who to test first within each risk group (newly symptomatic (solid lines) or first-symptomatic (dashed lines)). The results are stratified for three different levels of additional contact reduction due to symptoms (subplots) as well as for varying levels of additional contact reduction of high-risk individuals (x-axis). Linear regression fits are shown.

**Figure 5:**
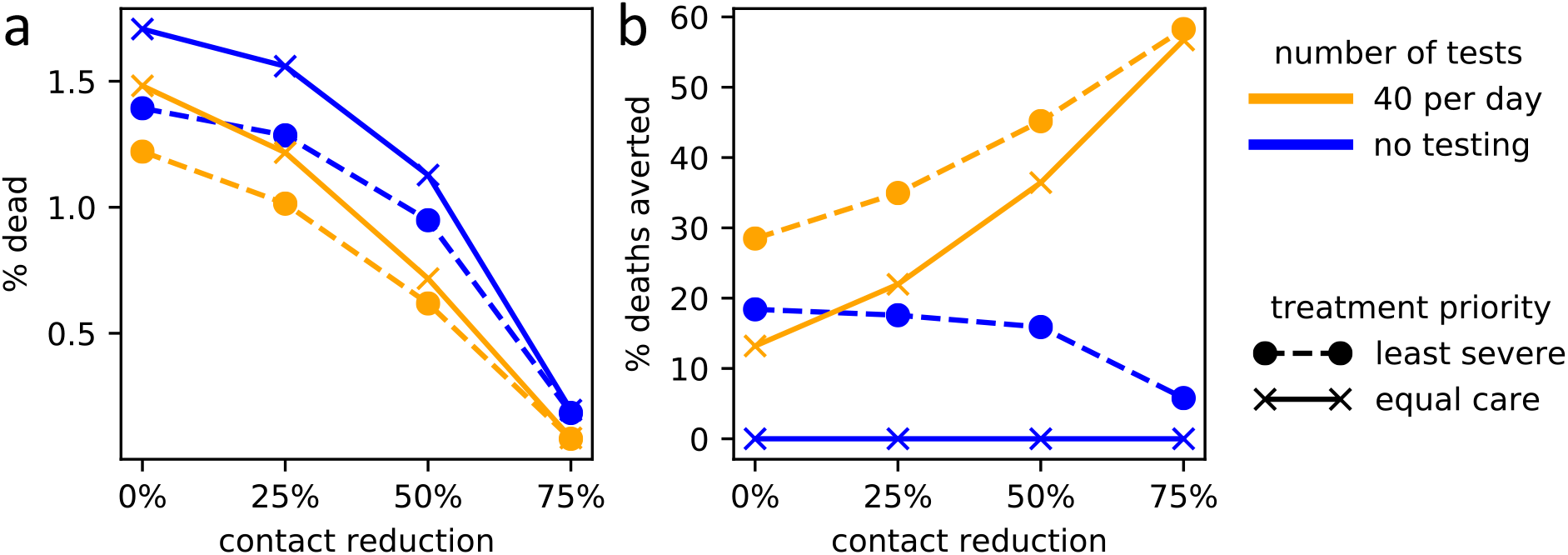
Interactions of the three policy domains on COVID-19-related deaths. The level of social contact reduction (x-axis) is plotted against combinations of testing policies (solid lines, gold: maximum testing, blue: no testing) and triage policies (dashed lines, circles: treat least severely infected first; solid lines, crosses: imperfect but equal care). (a) Policy choices versus proportion dead in the community. (b) Relative effects of policy choices in reducing the proportion dead, compared to the worst-case scenario of no testing and imperfect-but-equal-care triage policy.

### 2.4 Model outcomes

The primary model outcome considered in this study is the average number of deaths, or relatively speaking, the proportion of the population that dies from COVID-19. Related model outcomes, considered in Figs. S1, S2, S3, include the proportion of the population infected with SARS-CoV-2, the COVID-19 infection fatality rate (%dead / %infected), the COVID-19 case fatality rate (%dead / %symptomatic), the peak hospital (over)capacity (peak %hospitalized / capacity threshold), the initial basic reproductive number (average number of secondary infections caused by the individual who initially imports SARS-CoV-2 into the community), the time at which half of all infections happened (a measure of “flattening the curve” [53]), the average disease generation time (the time between infection of an individual and the time when the infecting person was infected), and the proportion of transmissions caused by asymptomatic cases (in compartment E or A).

### 2.5 Quantitative analysis

The model was implemented and all model analyses were run entirely in Python 3.7 [54]. The countour plots in Figs. 2, 3a, S5 were generated by binning the data using a 20×20 equidistant grid, and subsequent smoothing using a 2-dimensional Savitzky-Golay filter [55, 56]. To avoid over-smoothing, we chose a small window size of 5 and used only linear functions. Similarly, we used a one-dimensional Savitzky-Golay filter with window size 200 and linear functions to serve as a generalized moving average of the 1000 data points presented in Fig. 3b. In Figs. 4b,c, we summarized the raw data using a linear regression line.

## 3 Results

All model outcomes we investigated (Fig. S1,S2) were highly correlated (Fig. S3), therefore our results focus mainly on the proportion dead. Variation of virus- and disease-related parameters across the parameter space (Table S1) yielded an average initial basic reproductive number (*R*_0_) of 2.76 and an average disease generation time of 5.29 days (Fig. S2), both within the range of current estimates[3, 7, 48]. Higher transmissibility and higher *R*_0_ values were associated with faster generation times, which in turn were associated with hospital overcapacity and a faster spread of the virus (Fig. S4). Asymptomatic cases (truly asymptomatic or not yet symptomatic) caused most infections, which explains the ease with which SARS-CoV-19 is spreading across the world (Fig. S5). Interestingly, the average contact reduction of symptomatic cases influenced the proportion of infections caused by asymptomatic cases more than the rate of actual asymptomatic cases.

As expected, social distancing measures reduced the number of deaths. When we considered interaction networks with the same number of public and private contacts, a reduction in public contacts had a stronger effect on the number of deaths than an equal reduction in private contacts (Figure 2). The behavioral response of symptomatic individuals influences the level of overall contact reduction needed to keep hospitals under their capacity threshold (Figure 3a). However, hospitals quickly reach and surpass their capacity unless very strong levels of overall social distancing are implemented, even assuming perfect isolation of symptomatic individuals.

We compared different triage policies found in the literature to a worst-case scenario in which hospitals operating at overcapacity provide an imperfect but equal level of care to all patients (Fig. 3b) [12, 13, 16]. Current hospital policy of providing limited resources to the least-severely infected patients (based on clinical judgment) proved the most successful in reducing the proportion dead. Of the policies we investigated, the next most successful was to fill empty beds on a first-come first-served basis, while a policy which fills empty beds randomly resulted in a very similar number of deaths as imperfect-but-equal-care.

Increasing the availability of testing and reducing the delay between administering a test and receiving results both reduce the total number of deaths (Fig. 4a). We investigated the effect of a reduction in social activity by infected individuals who have tested positive under efficient and inefficient testing scenarios. In an efficient testing scenario, 20 to 40 symptomatic individuals (2 *−* 4% of the community) are tested per day and there is at most a one day delay between test administration and results, while under inefficient testing only one or two individuals are tested per day and results take 6 to 7 days. Our model indicates that effective quarantining of positively tested individuals (i.e., increased contact reduction) results in a meaningful reduction in deaths when testing is efficient, while it proved to have only a small effect when testing is inefficient (Fig. 4b).

Our model only considers the effect of testing on social behavior and not clinical outcomes, but within this framework we found that prioritizing testing of low-risk individuals consistently reduces the number of deaths more than testing high-risk individuals first (Fig. 4c). When we analyzed the difference between testing recently-infected individuals first versus prioritizing individuals who have been infected longer, we found that testing recently infected individuals first is more effective.

Our results point to interaction effects among the three policy domains we investigated. Policies influence each other: the efficacy of a particular policy choice depends on what other policies are implemented (Fig. 5). When social distancing is low, the choice of triage policy makes the biggest difference to the number of deaths, more so than the choice of testing policy. When social distancing is high however, triaging choices hardly matter, while testing becomes proportionately more important.

## 4 Discussion

Consistent with other COVID-19 related modeling studies (e.g. [3, 7, 20, 21]), our model results generally support current policies to reduce the public health impacts of the SARS-CoV-2 virus and COVID-19 infection.

A possible refinement of current social distancing policies is suggested by our comparison of interaction networks. We found that, for equally connected networks, a reduction in public contacts has a stronger effect on the number of deaths than an equal reduction in private contacts (Figure 2). This may be because public encounters allow the virus to spread randomly throughout the entire community, while private contacts enable only local spread. Based on these findings, future public health notices could increase their emphasis on the need to reduce contact with random individuals in the community.

Considering interaction networks with different levels of social connection (Figure 2) may provide an explanation for differences in the impact of the virus in different countries. A seminal study conducted in eight European countries found average per-day contact numbers of about 8 in Germany, 14 on average, and 20 in Italy [9]. At the time of this writing, Italy has suffered devastating effects from COVID-19 while Germany has weathered the pandemic relatively well. Because of the cultural differences in social connectivity, if Germany and Italy (for example) both reduce their contacts by the same percentage, Germany will avert a higher percentage of deaths. Our results suggest that more social societies must reduce their contacts more in order to attain the same level of reduction, and that a potentially fruitful avenue for future research might investigate whether these results hold more broadly across different cultures.

Currently, CDC guidelines prioritize the testing of symptomatic high-risk over low-risk individuals. Their goal is to ensure that individuals with a higher risk of complications are identified early and appropriately triaged [52]. Our model does not account for heterogeneity in clinical outcomes due to testing; the only effect of testing is on behavior through the reduction in social activity as the result of a positive test. Under this framework, our results indicate that prioritizing the testing of low-risk individuals more effectively reduces the overall death count than prioritizing the testing of high-risk individuals. Low-risk individuals have higher contact rates than high-risk individuals [9], so reducing the social activity of infected low-risk individuals slows the spread of the disease. Recognizing these differences in social activity levels may be important to the CDC when considering testing guidelines, as there may exist a trade-off between a short-term reduction in deaths due to appropriate triage of high-risk individuals and a long-term, overall higher death count caused by this choice.

The importance of testing policy is further highlighted by our interaction analysis. Several of our results suggest that policies influence each other, so that the efficacy of a particular policy choice depends on what other policies are implemented. The strength of social distancing of symptomatic individuals affects whether symptomatics or asymptomatics drive disease dynamics (Fig. S5), as well as the level of overall contact reduction needed to keep hospitals below capacity (Fig. 3a). Examining all three policy domains together, the relative efficacy of testing and triaging depends on the level of social distancing (Fig. 5).

In order to keep the model both tractable and understandable, we considered a basic contact network that differentiates between private and public contacts as well as high-risk and low-risk individuals. We did not consider heterogeneity in connectivity or individual risk behavior (e.g., super-spreaders [26]), or adaptation of policies over time. These homogeneities may explain the bimodal posterior distribution of the proportion of infected individuals (Fig. S2). Future studies may reveal if and how our results might change in more realistic interaction networks [57], which may include temporal movement of individuals [58], age-assortative mixing [21, 30], or the presence of individual-based heterogeneity in risk behavior.

Many key parameters of the COVID-19 epidemic are still unknown and may vary from community to community. We therefore ran our stochastic transmission model for a large variety of parameter settings. Due to the inherent uncertainty in many model parameters however, the model-generated, absolute values of the response variables may not be explicitly meaningful, as they depend on the particularities of the underlying parameter space. The goal of our study is not to report model predictions such as the expected absolute number of deaths; rather, our model is a tool that can be used, despite the uncertainty in key parameters, to compare the efficacy of various policies aimed at reducing the societal impact of COVID-19. We have focused on relative comparisons of three policy domains, as relative findings are more robust to inaccuracies in the underlying parameter space.

The presented model considers a variety of important factors in the current COVID-19 pandemic, in-cluding high-risk versus low-risk groups, differing social distancing behaviors, and the uncertainty around the proportion and infectivity of asymptomatic cases. Importantly, the model can be easily expanded and updated as more details about SARS-CoV-2 and COVID-19 emerge. Our results support current policies to contain the outbreak and suggest possible refinements to public health policy and education. Our results also provide a possible explanation for why some societies are more successful at containing the outbreak despite the implementation of similar policy measures. Follow-ups to this study could deepen our understanding of how heterogeneity in network structure and risk behavior affect the interplay of policy decisions and disease dynamics.

## Data Availability

The complete Python implementation of the model is available at Github at https://github.com/ckadelka/COVID19-network-model.

## Acknowledgement

We thank Rana Parshad and Zhijun Wu for fruitful initial discussions, Bernard Lidicky and Miles Aronnax for help with high performance computing, Katharina Kusejko, Nancy Boury, Philip Dixon, and Carolyn Seyler for helpful conversations, comments, and clarifications.

## References

(1) WHO 2020, url = https://www.who.int/emergencies/diseases/novel-coronavirus-2019/events-as-they-happen (accessed 19 April 2020).

(2) CDC 2020, url=https://www.cdc.gov/flu/pandemic-resources/1918-pandemic-h1n1.html accessed (13 April 2020).

(3) Ferguson, N., Laydon, D.; Nedjati Gilani, G.; Imai, N.; Ainslie, K.; Baguelin, M.; Bhatia, S.; Boonyasiri, A.; Cucunuba Perez, Z.; Cuomo-Dannenburg, G.; et al. 2020, DOI: https://doi.org/10.2556177482.

(4) Hellewell, J.; Abbott, S.; Gimma, A.; Bosse, N. I.; Jarvis, C. I.; Russell, T. W.; Munday, J. D.; Kucharski, A. J.; Edmunds, W. J.; Sun, F.; et al. The Lancet Global Health 2020.

(5) Funk, S.; Salathé, M.; Jansen, V. A. Journal of the Royal Society Interface 2010, 7, 1247–1256.

(6) Wilder-Smith, A.; Freedman, D. Journal of travel medicine 2020, 27, taaa020.

(7) Sanche, S.; Lin, Y. T.; Xu, C.; Romero-Severson, E.; Hengartner, N. W.; Ke, R. medRxiv 2020.02.07.20021154 (11 Feb 2020) 2020.

(8) Eksin, C.; Shamma, J. S.; Weitz, J. S. Scientific reports 2017, 7, 44122.

(9) Mossong, J.; Hens, N.; Jit, M.; Beutels, P.; Auranen, K.; Mikolajczyk, R.; Massari, M.; Salmaso, S.; Tomba, G. S.; Wallinga, J.; et al. PLoS medicine 2008, 5.

(10) Greenstone, M.; Nigam, V. University of Chicago, Becker Friedman Institute for Economics Working Paper No. 2020-26, url https://ssrn.com/abstract=3561244 accessed (13 April 2020).

(11) Kissler, S. M.; Tedijanto, C.; Goldstein, E.; Grad, Y. H.; Lipsitch, M. Science 2020.

(12) Emanuel, E. J.; Persad, G.; Upshur, R.; Thome, B.; Parker, M.; Glickman, A.; Zhang, C.; Boyle, C.; Smith, M.; Phillips, J. P. New Englad Journal of Medicine 2020.

(13) Biddison, E. L. D.; Faden, R.; Gwon, H. S.; Mareiniss, D. P.; Regenberg, A. C.; Schoch-Spana, M.; Schwartz, J.; Toner, E. S. Chest 2019, 155, 848–854.

(14) Zucker, H. A. Ventilator Allocation Guidelines; tech. rep., New York State Department of Health, 2015.

(15) Kanter, R. K. Chest 2015, 147, 102–108.

(16) Powell, T.; Christ, K. C.; Birkhead, G. S. Disaster Medicine and Public Health Preparedness 2008, 2, 20–26.

(17) Sharfstein, J. M.; Becker, S. J.; Mello, M. M. JAMA 2020.

(18) Goldstein, N. D.; Burstyn, I. 2020, DOI: doi:10.31219/osf.io/9pz4d..

(19) CDC Evaluating and testing persons for coronavirus disease 2019 (COVID-19), url = https://www.cdc.gov/coronavincov/hcp/clinical-criteria.html accessed (20 April 2020), 2020.

(20) Kretzschmar, M.; Rozhnova, G.; van Boven, M. 2020.

(21) Prem, K.; Liu, Y.; Russell, T. W.; Kucharski, A. J.; Eggo, R. M.; Davies, N.; Flasche, S.; Cli↵ord, S.; Pearson, C. A.; Munday, J. D.; et al. The Lancet Public Health 2020.

(22) Eberhardt, J. N.; Breuckmann, N. P.; Eberhardt, C. S. medRxiv 2020.04.10.20061176 14 April 2020 2020.

(23) Rodriguez, P. F. medRxiv 2020.04.01.20050393 6 April 2020 2020.

(24) Meyers, L. A.; Pourbohloul, B.; Newman, M. E.; Skowronski, D. M.; Brunham, R. C. Journal of theoretical biology 2005, 232, 71–81.

(25) Bansal, S.; Grenfell, B. T.; Meyers, L. A. Journal of the Royal Society Interface 2007, 4, 879–891.

(26) Lloyd-Smith, J. O.; Schreiber, S. J.; Kopp, P. E.; Getz, W. M. Nature 2005, 438, 355–359.

(27) Pastor-Satorras, R.; Castellano, C.; Van Mieghem, P.; Vespignani, A. Reviews of modern physics 2015,87, 925.

(28) Guan, W.-j.; Ni, Z.-y.; Hu, Y.; Liang, W.-h.; Ou, C.-q.; He, J.-x.; Liu, L.; Shan, H.; Lei, C.-l.; Hui, D. S., et al. New England Journal of Medicine 2020.

(29) Bi, Q.; Wu, Y.; Mei, S.; Ye, C.; Zou, X.; Zhang, Z.; Liu, X.; Wei, L.; Truelove, S. A.; Zhang, T.; et al.medRxiv 2020.03.03.20028423 (27 March 2020) 2020.

(30) Miller, J. C.; Volz, E. M. PloS One 2013, 8.

(31) Anderson, R. M.; Heesterbeek, H.; Klinkenberg, D.; Hollingsworth, T. D. The Lancet 2020, 395, 931–934.

(32) Watts, D. J.; Strogatz, S. H. Nature 1998, 393, 440.

(33) Liu, Q.-H.; Ajelli, M.; Aleta, A.; Merler, S.; Moreno, Y.; Vespignani, A. Proceedings of the National Academy of Sciences 2018, 115, 12680–12685.

(34) CDC National Center for Immunization and Respiratory Diseases (NCIRD) Division of Viral Diseases 2020, url=https://www.cdc.gov/coronavirus/2019-ncov/specific-groups/high-risk-complications.html accessed (19 April 2020).

(35) WHO Consensus document on the epidemiology of severe acute respiratory syndrome (SARS); tech. rep., World Health Organization, 2003.

(36) Yu, P.; Zhu, J.; Zhang, Z.; Han, Y.; Huang, L. The Journal of infectious diseases 2020.

(37) Mizumoto, K.; Kagaya, K.; Zarebski, A.; Chowell, G. Eurosurveillance 2020, 25, 2000180.

(38) Rothe, C.; Schunk, M.; Sothmann, P.; Bretzel, G.; Froeschl, G.; Wallrauch, C.; Zimmer, T.; Thiel, V.; Janke, C.; Guggemos, W.; et al. New England Journal of Medicine 2020.

(39) Fraser, C.; Riley, S.; Anderson, R. M.; Ferguson, N. M. Proceedings of the National Academy of Sciences 2004, 101, 6146–6151.

(40) Woelfel, R.; Corman, V. M.; Guggemos, W.; Seilmaier, M.; Zange, S.; Mueller, M. A.; Niemeyer, D.; Vollmar, P.; Rothe, C.; Hoelscher, M.; et al. medRxiv 2020.03.05.20030502 (8 March 2020) 2020.

(41) Wu, Z.; McGoogan, J. M. Jama 2020.

(42) Verity, R.; Okell, L. C.; Dorigatti, I.; Winskill, P.; Whittaker, C.; Imai, N.; Cuomo-Dannenburg, G.; Thompson, H.; Walker, P. G.; Fu, H.; et al. The Lancet Infectious Diseases 2020.

(43) Koma, W.; Neuman, T.; Claxton, G.; Rae, M.; Kates, J.; Michaud, J. 2020, url=https://www.k↵.org/global-health-policy/issue-brief/how-many-adults-are-at-risk-of-serious-illness-if-infected-with-coronavirus/ accessed (13 April 2020).

(44) Bureau, U. C. 2017, url=https://www.census.gov/data/datasets/2017/demo/popproj/2017-popproj.html accessed 18 April 2020.

(45) Lauer, S. A.; Grantz, K. H.; Bi, Q.; Jones, F. K.; Zheng, Q.; Meredith, H. R.; Azman, A. S.; Reich, N. G., Lessler, J. Annals of Internal Medicine 2020.

(46) Zhang, J.; Litvinova, M.; Wang, W.; Wang, Y.; Deng, X.; Chen, X.; Li, M.; Zheng, W.; Yi, L.; Chen, X.; et al. The Lancet Infectious Diseases 2020.

(47) Zhou, F.; Yu, T.; Du, R.; Fan, G.; Liu, Y.; Liu, Z.; Xiang, J.; Wang, Y.; Song, B.; Gu, X.; et al. The Lancet 2020.

(48) Wang, D.; Hu, B.; Hu, C.; Zhu, F.; Liu, X.; Zhang, J.; Wang, B.; Xiang, H.; Cheng, Z.; Xiong, Y.; et al. Jama 2020.

(49) Huang, C.; Wang, Y.; Li, X.; Ren, L.; Zhao, J.; Hu, Y.; Zhang, L.; Fan, G.; Xu, J.; Gu, X.; et al. The Lancet 2020, 395, 497–506.

(50) Pan, F.; Ye, T.; Sun, P.; Gui, S.; Liang, B.; Li, L.; Zheng, D.; Wang, J.; Hesketh, R. L.; Yang, L.; et al. Radiology 2020, 200370.

(51) OECD Health at a Glance 2019: OECD Indicators; tech. rep., Organisation for Economic Co-operation and Development, 2019.

(52) CDC 2020, url=https://www.cdc.gov/coronavirus/2019-ncov/cases-updates/testing-in-us.html accessed (19 April 2020).

(53) Qualls, N.; Levitt, A.; Kanade, N.; Wright-Jegede, N.; Dopson, S.; Biggersta↵, M.; Reed, C.; Uzicanin, A.; Group, C. C. M. G. W.; Group, C. C. M. G. W.; et al. MMWR Recommendations and Reports 2017, 66, 1.

(54) Team, P. C. Python: A dynamic, open source programming language; Python Software Foundation, 2020.

(55) Savitzky, A.; Golay, M. J. Analytical chemistry 1964, 36, 1627–1639.

(56) Pan, B.; Xie, H.; Guo, Z.; Hua, T. Optical Engineering 2007, 46, 033601.

(57) Salathé, M.; Kazandjieva, M.; Lee, J. W.; Levis, P.; Feldman, M. W.; Jones, J. H. Proceedings of the National Academy of Sciences 2010, 107, 22020–22025.

(58) Eubank, S.; Guclu, H.; Kumar, V. A.; Marathe, M. V.; Srinivasan, A.; Toroczkai, Z.; Wang, N. Nature 2004, 429, 180–184.

